# Role of common and rare genetic variants in the aetiology of trigeminal neuralgia

**DOI:** 10.1101/2024.07.16.24310509

**Authors:** Kim J. Burchiel, Olga A. Korczeniewska, Fengshen Kuo, Ching-Yu Huang, Ze’ev Seltzer, Scott R Diehl

## Abstract

**Background:** Trigeminal neuralgia (TN) is characterized by repeated paroxysmal attacks of severe facial pain usually lasting 1-3 minutes. Lifetime prevalence is ca.3 per 1,000, more common in women, and with onset generally in middle age. Medications usually provide relief in the early stages of the disorder, but for many patients, severe drug side effects emerge and medically intractable pain returns, sometimes lasting for life. Some patients present with paroxysmal pain predominantly while others also experience substantial concomitant constant facial pain. Some patients have a history of a blood vessel compressing and damaging their trigeminal nerve (neurovascular compression, NVC). For these “classical” cases, surgery often provides complete or substantial pain relief for many years. “Idiopathic” cases without NVC or any other apparent cause also occur. NVC was previously observed to be less frequent in females who had early age of onset and these patients may constitute a unique subgroup. Our aim was to evaluate the role of inherited genetic variation in the aetiology of TN in patient subgroups based on age of onset, presence of NVC and sex.

**Methods:** To maximize aetiological homogeneity, only patients with predominantly paroxysmal pain and minimal concomitant continuous pain were included in the analysis. Conditions known to cause secondary TN such as tumors or multiple sclerosis were excluded. The GWAS analysis was based on 626 TN patients and 827 Control subjects of European ancestry recruited in Canada, the UK, and US. A Genome-Wide Association Study (GWAS) analysis was performed using Affymetrix’s Precision Medicine arrays yielding 7,781,254 biallelic DNA variants available after Quality Control (QC) and imputation. Rare damaging mutations in genes with functions relevant to the biology of TN were identified in Whole Genome Sequencing (WGS) genomic DNA of 100 patients using a novel strategy based on overlap of symptoms of TN with symptoms of known genetic disorders.

**Findings:** The GWAS analysis revealed associations at eight genome locations including near *LRP1B* (P-value 6.3 X 10^-15^), a gene important for repair of myelin sheath injury that has been previously proposed as a target for the treatment of neuropathic pain. Associations were also found for the potassium channel gene *KCNK10*, and for *CHL1, CUX1, SGMS1* and *ZNF804B* genes, all genes with neural functions potentially relevant to the aetiology of TN. In addition, high-risk genotypes at the *CUX1* and *KCNK10* genes exhibit significant interactions with patients’ sex and the presence or absence of NVC (P-values 0.005 and 0.017, respectively). Whole genome sequencing of 100 TN patients revealed mutations in ion channel genes *TRPM4* (six patients), *SCN10A* and *SCNN1B* (five patients), *CACNA1F, CACNA1S* and SCN5A (four patients) and *CACNA1H*, *SCN2A* and *SCN9A* (three patients). Female patients with onset prior to age 46 had more mutated genes with myelin-related functions (P-value 0.004) and associated with epilepsy or seizure (P-value 0.03) than older onset females and males of any onset age.

**Interpretation:** Risk of TN in patients presenting with paroxysmal pain only is associated with both common genetic variants and with rare mutations. Some high-risk genotypes have significant interactions with sex and NVC. Evidence of the condition’s heterogeneous genetic aetiology should be considered when evaluating novel therapies.

**Funding:** Grants from the William H. and Leila A. Cilker Genetics Research Program of the Facial Pain Research Foundation, The Foundation of the University of Medicine and Dentistry of New Jersey, and Rutgers School of Dental Medicine, Rutgers Health, Rutgers – The State University of New Jersey

**Contact:** Scott R Diehl, PhD, 973-972-7053

## Introduction

All TN cases are characterized by repeated paroxysms of severe lancinating facial pain lasting 1-3 minutes often described as “electrical” in quality. While some patients experience predominantly paroxysmal pain (TN1) others suffer from substantial constant pain between paroxysms (TN2) (Burchiel 2003). The International Classification of Headache Disorders (ICHD-3)(Olesen 2024) further subdivides TN into “Idiopathic” cases (lacking neurovascular compression, NVC) and “Classical” cases (those with “demonstration on MRI or during surgery of neurovascular compression (not simply contact), with morphological changes in the trigeminal nerve root.” TN1 cases can be Classical or Idiopathic and the same is true for TN2, so there are four subtypes identified by this classification scheme. Under the assumption that it may yield a more genetically homogeneous case group, recruitment for this study was limited to patients with pain that is predominantly paroxysmal (TN1), including both Classical (ICHD-3 13.1.1.1.1) and Idiopathic (13.1.1.3.1) cases.

Lifetime prevalence of TN is ca. 3 per 1,000, more common in women (Ashina et al., 2024). Onset is usually in middle age but female cases with an early age of onset have previously reported to be more likely to lack NVC and may represent a unique patient subgroup (Ko et al., 2015). Medications usually provide relief in the early stages of the disorder, but adverse side effects often develop, and medically intractable pain can reemerge. TN frequently has devastating psychological, social, and economic impacts on patients’ lives (Bendtsen et al., 2020). For cases of classical TN with severe NVC, microvascular decompression surgery often provides complete or substantial pain relief for many years, although pain sometimes returns. Development of more effective treatments is likely to be accelerated by an improved understanding of the molecular and physiological processes that underlie the development of TN including genetic differences among individual patients. Limited studies of aggregation of TN cases in families and tests of candidate polymorphisms have been previously reported (Panchagnula et al., 2019; DiStefano et al., 2020; Eide 2022; Wang et al., 2023) but no highly penetrant dominant genes have been found with large numbers of cases occurring across multiple generations. The limited number of GWAS and DNA sequencing studies reported to date have thus far failed to reveal strong genetic associations of either common polymorphisms or rare mutations that have been independently replicated (Ashina et al., 2024; Dong et al., 2023). The aim of this study was to develop a well-characterized homogeneous group of TN cases to assess the role of inherited genetic variation in the aetiology of TN, and to consider potential effects attributable to patients’ age of TN onset, sex, and presence or absence of NVC.

## Methods

### Study design and participants

The GWAS sample included 626 TN patients (65% female) of European ancestry recruited at nine locations in Canada, the UK, and US (details in Supplementary Table S1) with human subjects IRB (ethics) review of protocols approved at each site. A Control group had 827 subjects (57% female) also of European ancestry obtained from previous studies unrelated to any form of pain or neurological disorder. Age of TN onset averaged 54.9 (15 – 91) years in females and 56.1 (17 – 89) years in males. NCV data were available for 294 TN1 cases. Females with age of TN onset <=45 versus >45 years were classified as idiopathic 22% and 30% of the time, respectively, and males were idiopathic 20% and 17%, respectively, in these age of onset groups. Patients who report experiencing a limited amount of concomitant constant facial pain were eligible for recruitment if their paroxysmal pain clearly predominated (TN1)(Burchiel 2003). To maximize aetiological homogeneity, patients with substantial concomitant continuous pain (TN2) or conditions known to cause secondary TN such as tumors or multiple sclerosis were excluded. Patients were classified as TN1 by clinicians with substantial experience diagnosing TN. The study’s cases most closely match the IHS ICHD-3 13.1.1.1, “classical trigeminal neuralgia, purely paroxysmal or 13.1.1.3.1, “idiopathic trigeminal neuralgia, purely paroxysmal diagnoses, except that our TN1 definition only requires that their paroxysmal pain be predominant but not necessarily “purely paroxysmal.” In addition, to be eligible patients also had to be diagnosed as TN1 using a neural network-based questionnaire (McCartney et al., 2014). Genetic data revealed previously unreported pairs of relatives among TN1 cases (a pair of siblings and a parent-offspring duo). Saliva was obtained from subjects using Oragene•DISCOVER saliva collection kits (DNA Genotek, Ottawa, Canada).

### Procedures

DNA was extracted from saliva using methods recommended by the collection kit manufacturer. SNPs and small insertions and deletions (in/dels) for the GWAS were assayed using Affymetrix Precision Medicine Research Array and genotypes were called using the Axiom Analysis Suite software (Thermo Fisher Scientific Inc., Waltham, MA). Whole Genome DNA Sequencing (WGS, PCR-free 30X coverage) was performed for 100 TN cases (a subset of GWAS cases with highest quality DNA) at the Broad Clinical Labs (subsidiary of Broad MIT and Harvard University, Boston, MA). CRAM and VCF files of SNPs and small in/del mutations for each patient were generated using the Broad Clinical Lab’s GATK informatics pipeline (DePristo et. al., 2017).

### Statistical analysis

GWAS analyses of SNPs and in/dels were performed using the SNP & Variation Suite software package of Golden Helix, Inc. (Bozeman, MT). After QC and imputation using the Beagle program (Browning et al., 2018) a total of 7,781,25 biallelic DNA variants were available for the GWAS analysis. Principal Components Analysis (PCA) was used to exclude samples falling outside of the distinct cluster of 626 TN Cases and 827 Controls of European ancestry. A single locus mixed model (Segura et al., 2012) with additive transmission and with sex as a covariate was used for tests of association for the GWAS data. Q-Q plot analysis shows that the single locus mixed model robustly adjusted for potential P-value inflation due to cryptic relatedness or other violations of model assumptions (Supplementary Figure S1). High-Risk Genotypes (HRGs) at associated SNPs were defined as homozygous and/or heterozygous genotypes with higher frequency in Cases than Controls, and Low-Risk Genotypes (LRGs) were defined as the genotypes at lower frequency in Cases than Controls. The Genomic Best Linear Unbiased Prediction (GBLUP) method (Wang et al., 2018) was used to assess how accurately the eight most strongly associated SNPs predict Case versus Control status of subjects included in the GWAS. Area Under the Curve (AUC) using the Wilcoxon Mann Whitney method and Sensitivity and Specificity were estimated. Lastly, interactions between SNP HRGs, sex and idiopathic versus classical TN subtypes were evaluated.

Rare mutations predicted to alter (usually damage) protein function in genes with functions relevant to the biology of TN were identified in Whole Genome DNA Sequencing (WGS) data of 100 TN patients using a novel strategy based, in part, on similarity of symptoms of TN patients with those of known genetic disorders, as described below. This analysis was performed using Fabric Genomics (Oakland, CA) proprietary software that ranks the annotated variants using the VAAST (Flygare et al., 2018) and Phevor (Singleton et al., 2014) programs. WGS data were also analyzed using Emedgene (Illumina, Inc.) software that uses a similar approach (Meng et al., 2023). The method requires input of the signs and symptoms of the disorder under study using standardized vocabulary of the Human Phenotype Ontology (HPO) database for phenotypic abnormalities observed in human diseases (Köhler et al., 2017). Analyses were performed using three overlapping sets of HPO terms for TN, ranging from very narrow (only the term “Trigeminal neuralgia (HP: 0100661) to longer lists of 12 and 31 signs and symptoms relevant to TN (Supplementary Table S2). Both the Fabric Genomics and the Emedgene approaches produce lists of top candidate variants for each TN Case ranked based on Minor Allele Frequency (MAF, assumed rare, consistent with the low prevalence of TN), variants predicted to cause changes in protein function (most often damaging) and evolutionary conservation. These algorithms performed complex searches for human disorders that have multiple signs and symptoms overlapping with the HPO terms selected for TN. The priority ranking of mutations found in TN patients is increased when the programs identify disorders with similar symptoms that have mutations in the same gene as the TN patient. These programs analyze one patient at a time so for this cohort study a custom merged file was created combining the prioritized variants for all 100 TN Cases. Finally, genes with high priority mutations in TN patients were ranked based on the extent to which their biological functions and associated disorders match the suspected pathophysiology of TN using information in the GeneCards database (Stelzer et al., 2016) and review of the literature using PubMed and Google searches. A comprehensive comparison of results obtained from the Fabric Genomics and Emedgene programs is beyond the scope of this paper, but in general they prioritized most of the same genes and mutations. Finally, each gene included on the final list of candidate mutations was classified as belonging to one or more of the following six functional-clinical categories: ion channels, myelin/Charcot-Marie Tooth disease, epilepsy/seizure, neuronal development, other nerve functions and/or pain.

## Results

### Genome Wide Association Study (GWAS)

Associations with genome-wide statistical significance (P-value < 5 X10^-8^) and suggestive evidence of association (P-value < 1 X 10^-6^) found in eight regions of the genome are shown in Figure 1. SNPs with the smallest P-values in each region and nearest genes are shown in Table 1. Genome wide statistical significance was found in four regions including 145 kbp 3’ of the *LRP1B* gene (P-value = 6.3 X 10^-15^) and in an intron of the *CUX1* gene (P-value = 4 X 10^-8^).

**Figure 1:**
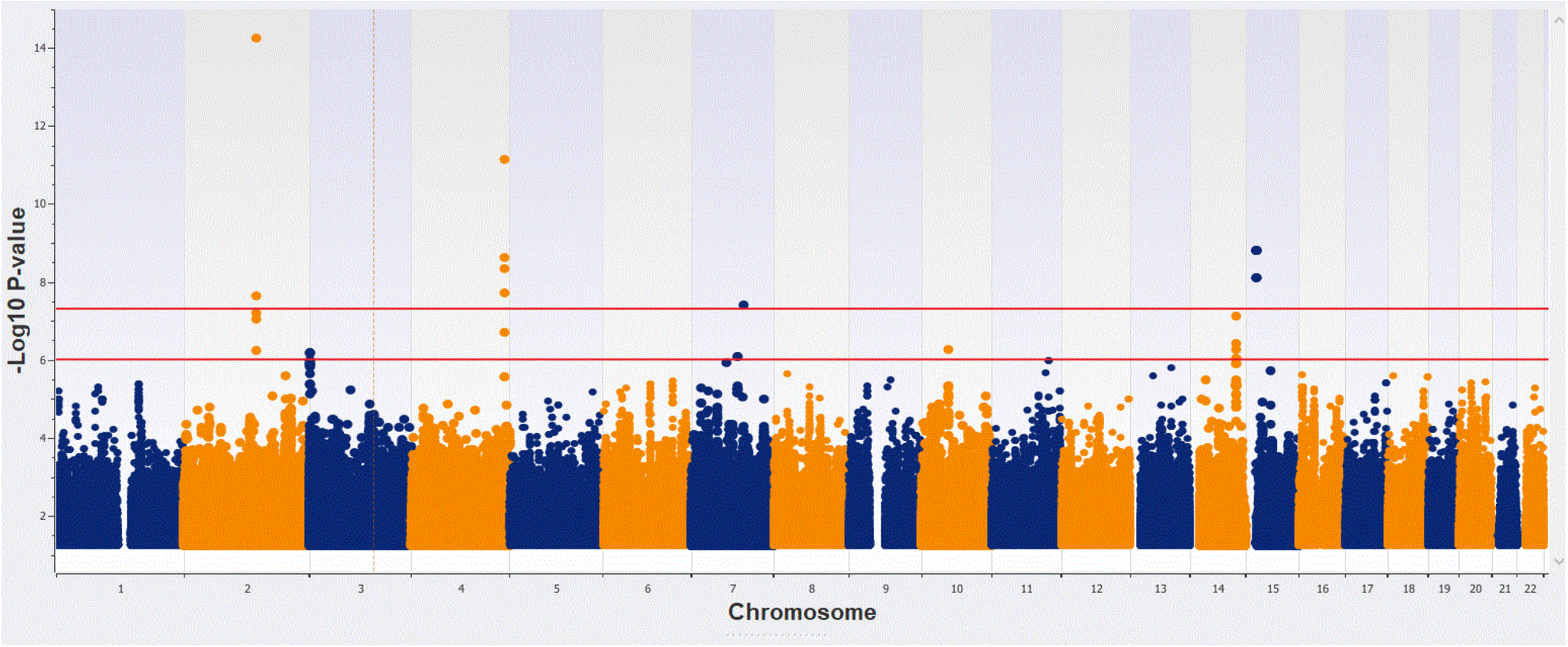
Manhattan plot of Genome Wide Association Study Results in TN patients of European Ancestry. The upper red horizontal line demarks the threshold of genome-wide statistical significance (-log10 P-value > 7.3) and the lower line demarks “suggestive” evidence of association (-log10 P-value > 6.0).

**Table 1:** Genetic polymorphisms associated with Trigeminal Neuralgia. GWAS associations discovered using a sex-adjusted Single Locus Mixed Model (SLMM) assuming additive transmission of risk.

| Marker | Chromosome | Position (hg19) | P-value | Nearest Gene (Symbol) | Position in/near Gene | Nearest Gene (Name) |
| --- | --- | --- | --- | --- | --- | --- |
| rs116640189 | 2 | 140,845,541 | $6.3 \times 10^{-15}$ | *LRP1B* | 145 kbp 3' | LDL Receptor Related Protein 1B |
| rs9819335 | 3 | 287,275 | $6.3 \times 10^{-7}$ | *CHL1* | Intron | Cell Adhesion Molecule L1 Like |
| rs75624142 | 4 | 181,396,367 | $7.9 \times 10^{-12}$ | *LINC00290* | 596 kbp 3' | Long Intergenic Non-Protein Coding RNA 290 |
| rs73390621 | 7 | 89,007,703 | $7.9 \times 10^{-7}$ | *ZNF804B* | 40 kbp 3' | Zinc Finger Protein 804B |
| rs4727512 | 7 | 101,558,899 | $4.0 \times 10^{-8}$ | *CUX1* | Intron | Cut Like Homeobox 1 |
| rs12217518 | 10 | 52,098,186 | $5.0 \times 10^{-7}$ | *SGMS1* | Intron | Sphingomyelin Synthase 1 |
| rs41286550 | 14 | 88,706,920 | $7.9 \times 10^{-8}$ | *KCNK10* | Intron | Potassium Two Pore Domain Channel Subfamily K Member 10 |
| rs572198006 | 15 | 20,086,440 | $1.6 \times 10^{-9}$ | *ENSG00000288375* | 39 kbp 3' | Protein coding gene of unknown function |

Genes closest to the two other regions with GWAS significance have unknown functions. In the four genome regions with suggestive evidence of association the top SNPs are in introns of the *KCNK10*, *CHL1*, *ZNF804B* and *SGMS1* genes. Frequencies of the HRGs of the eight top SNPs are shown in Figure 2 stratified by sex and age of onset. Six SNPs have very low HRG frequency in controls (0.1-1.9%) and moderately higher frequency in TN1 cases (4-12%). HRG frequencies of SNPs rs4727512 and rs12217518 in controls are much higher (30% and 76%, respectively) but higher still (41% and 84%, respectively) in TN1 cases. Cases carrying the HRG were more often idiopathic than cases carrying the LRG at four GWAS SNPs, about the same at two SNPs and less often idiopathic at two SNPs but none of these differences are statistically significant (Supplementary Figure S2).

**Figure 2:**
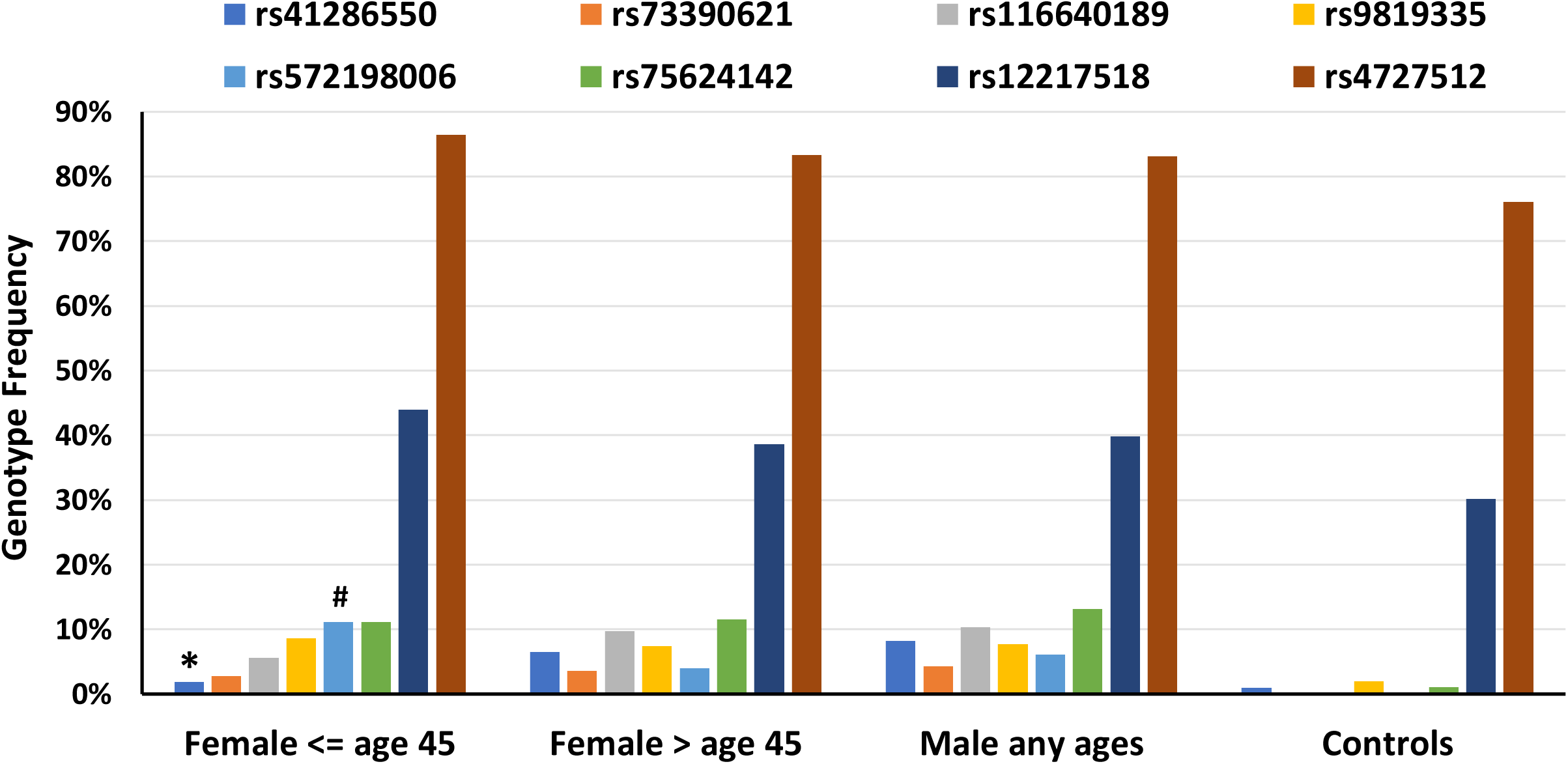
High-Risk Genotype (HRG) Frequency in TN Cases v. Controls. Frequency of Genotypes at higher frequency in TN Cases v. Controls at SNPs identified in the GWAS analysis shown in Figure 1 and Table 1. Female Cases with onset at age 45 or younger have lower HRG frequency at SNP rs41286550 (2%) than females with onset after age 45 (6%) and male Cases regardless of age of onset (8%) (*P-value 0.044). Female Cases with onset at age 45 or younger have higher HRG frequency at SNP rs572198006 (11%) than females with onset after age 45 (4%) and male Cases regardless of age of onset (6%) (^#^P-value = 0.018).

SNPs in Table 1 accurately predict TN1 case and control phenotype for subjects used in the GWAS with AUC 0.86, sensitivity 0.65 and specificity 0.85 (Figure 3). Evidence of three-way interactions of SNP genotypes, sex and idiopathic versus classical TN1 subtype was found for SNPs located in the *CUX1* (Figure 4A, P-value 0.005) and *KCNK10* genes (Figure 4B, P-value 0.017). Taking the interactions into account, idiopathic TN1 cases are more frequent in LRGs in both genes (P-values 0.004 and 0.043, respectively). Frequency of the *CUX1* HRG is higher in females than males (87% versus 82%, P-value 0.014) and idiopathic cases are also more frequent in females than males (28% versus 18%, P-value 0.006).

**Figure 3:**
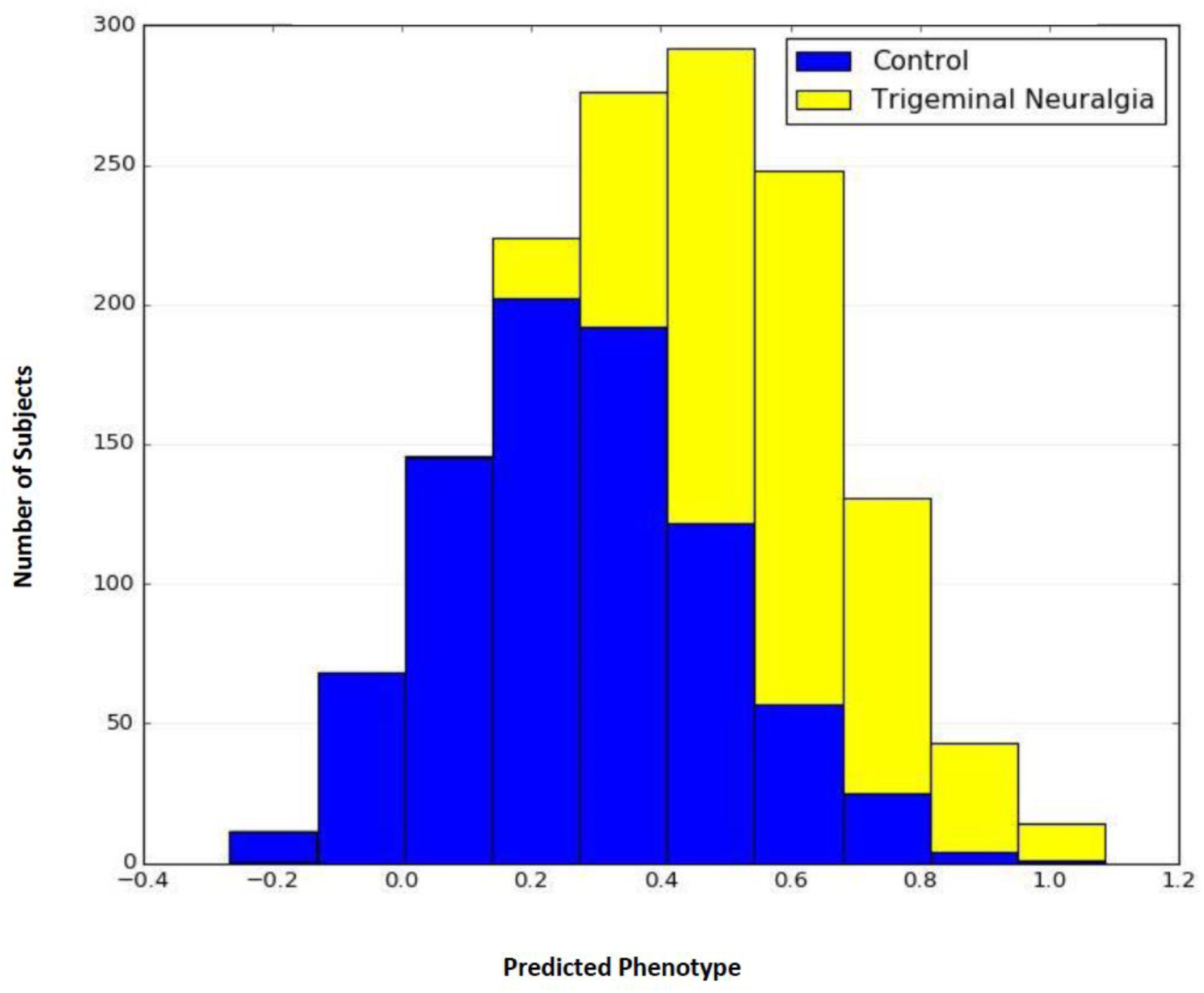
Genomic Best Linear Unbiased Prediction (GBLUP) of TN Cases v. Controls. Based on genotypes of 8 SNPs identified in the GWAS shown in Table 1. Additive genetic merits by sample and allele substitution effects by marker. Mean Predicted Phenotype score for 626 TN Cases is 0.57 with standard error 0.007 v. 0.28 with standard error 0.007 for 827 Controls (P-value 6.8E-145). Area Under the Curve (AUC, Using the Wilcoxon Mann Whitney method) is 0.86 with Sensitivity 0.65 and Specificity 0.85.

**Figure 4A.**
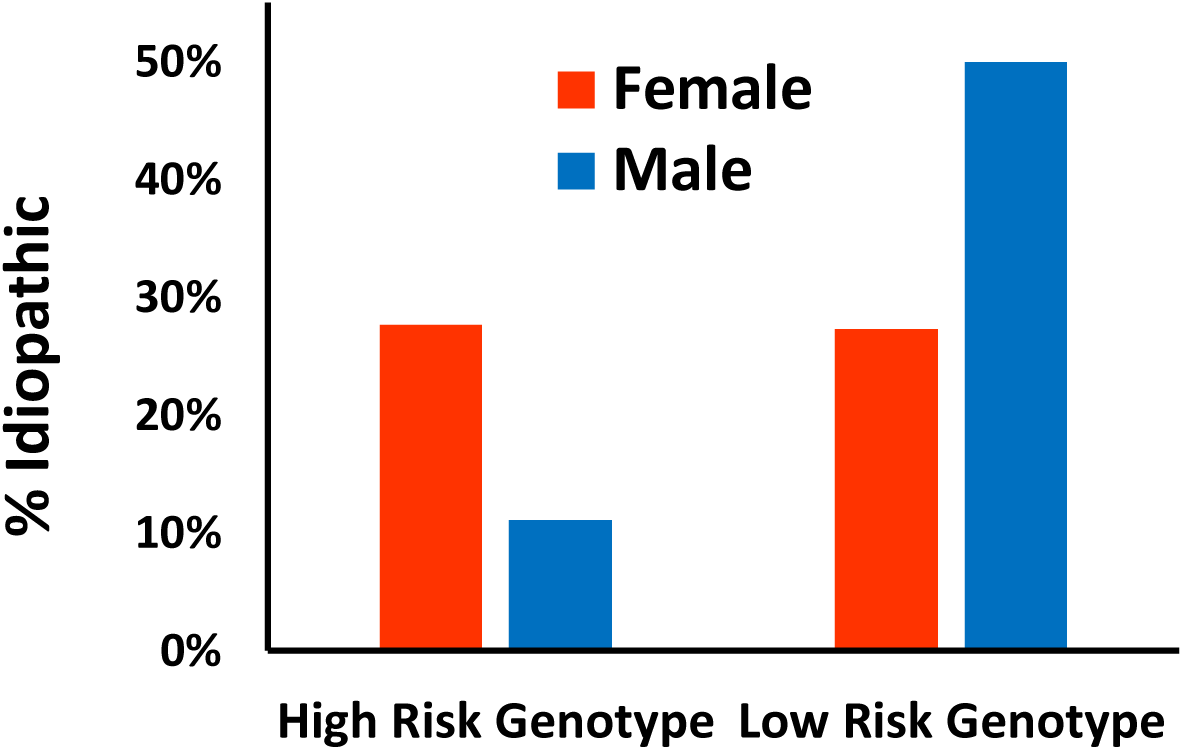
Interaction of *CUX1* Genotypes with Sex and Neurovascular Compression. Females are more frequently idiopathic than males when both are HRG at rs4727512 but idiopathic less often than males when both are LRG (p = 0.005); higher frequency of Idiopathic cases in LRG, (37%) v. HRG (22%)(p = 0.004); females more often HRG (87%) than Males (82%) (p = 0.014); and females more often Idiopathic (28%) than Males (18%) (p = 0.006).

**Figure 4B.:**
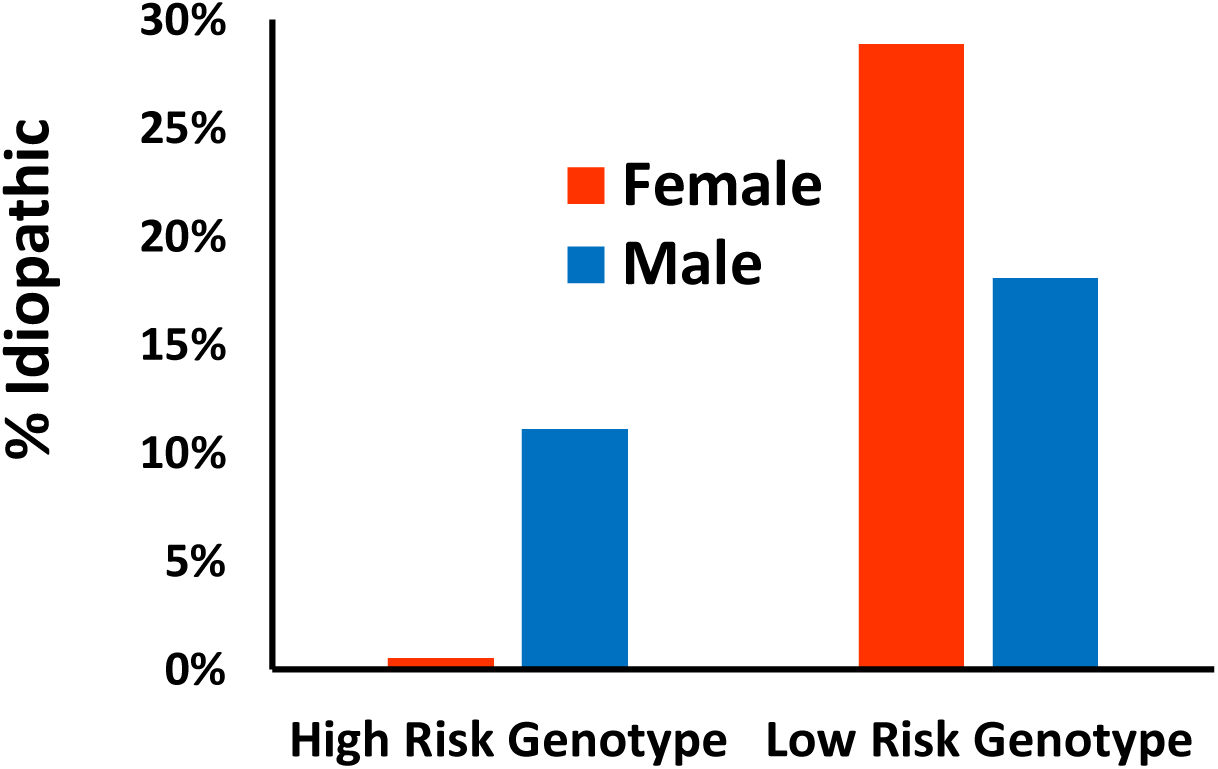
Interaction of *KCNK10* Genotypes with Sex and Neurovascular Compression. Females are less frequently idiopathic than males when both are HRG at rs41286550 but idiopathic more often than males when both are LRG (p-value 0.017); higher frequency of Idiopathic cases in LRG, (25%) v. HRG (5%)(p-value 0.043); No significant sex difference in HRG frequency of Females (5%) v. Males (10%) (p-value 0.172); females more often Idiopathic (28%) than Males (17%) (p-value 0.067).

### Whole Genome Sequencing (WGS)

A total of 349 mutations in 182 genes were identified in 100 TN patients sequenced including 92 genes with 2-8 mutations per gene and 73 genes with mutations found in 2-6 patients per gene (Supplementary Tables S3 and S4). All mutations were heterozygotes with the reference allele except for gene *TBC1D24*. Genes with mutations in 3-6 patients are show in Table 2 grouped according to their known biological functions with the largest numbers of genes having functions involving ion channels or in the myelin/Charcot-Marie-Tooth (CMT) disease group. Minor Allele Frequencies (MAF) of mutations reported in the gnomAD database (Chen et al., 2024) ranging from 0.00001 to 0.001 for 222 mutations, from 0.001 to 0.01 for 28 mutations and from 0.01 and 0.1 for 23 mutations. Mutations in the latter high MAF group were included as potential TN1 aetiology candidates only when they co-occurred in patients with another mutation of a lower allele frequency, thus potentially comprising a compound heterozygote of low genotype frequency. A total of 49 mutations are not reported in gnomAD and so their frequencies were unavailable, and 28 mutations had frequencies listed in gnomAD as 0.0000 for populations of European ancestry. The number of mutated genes per patient was significantly higher in female patients who experienced early onset of TN1 prior to age 46 compared to females with onset at older age or males regardless of onset age for genes with functions related to Myelin/CMT (P-value 0.005) or Epilepsy/Seizure activity (P-value 0.03) (Figure 5). Ten identical mutations were found in two patients. The one parent offspring pair among the study’s cases accounted for the co-occurrence of two of the pairs of identical mutations (in *CACNA1B* and *FIG4* genes). Four of the other pairs of identical mutations were of high enough MAF (0.01 to 0.06) to explain their co-occurrence among 100 unrelated individuals by chance. The remaining four pairs of identical mutations were in unrelated individuals, with MAF ranging from 0.0006 to 0.003, and are located in *DOCK3*, *NIPA1*, *SLC1A2* and *TRPM4* genes (Supplementary Table S3). Fabric Genomics’ Phevor scores used to rank mutations based on HPO terms labelled Models 2 and 3 (Supplementary Table S2) are highly correlated (r = 0.99). Both models are moderately correlated with Model 1 that includes only HPO term HP: 0100661 Trigeminal Neuralgia (r = 0.58 and 0.60) indicating that evaluations performed under different HPO models sometimes yield substantially different results.

**Figure 5:**
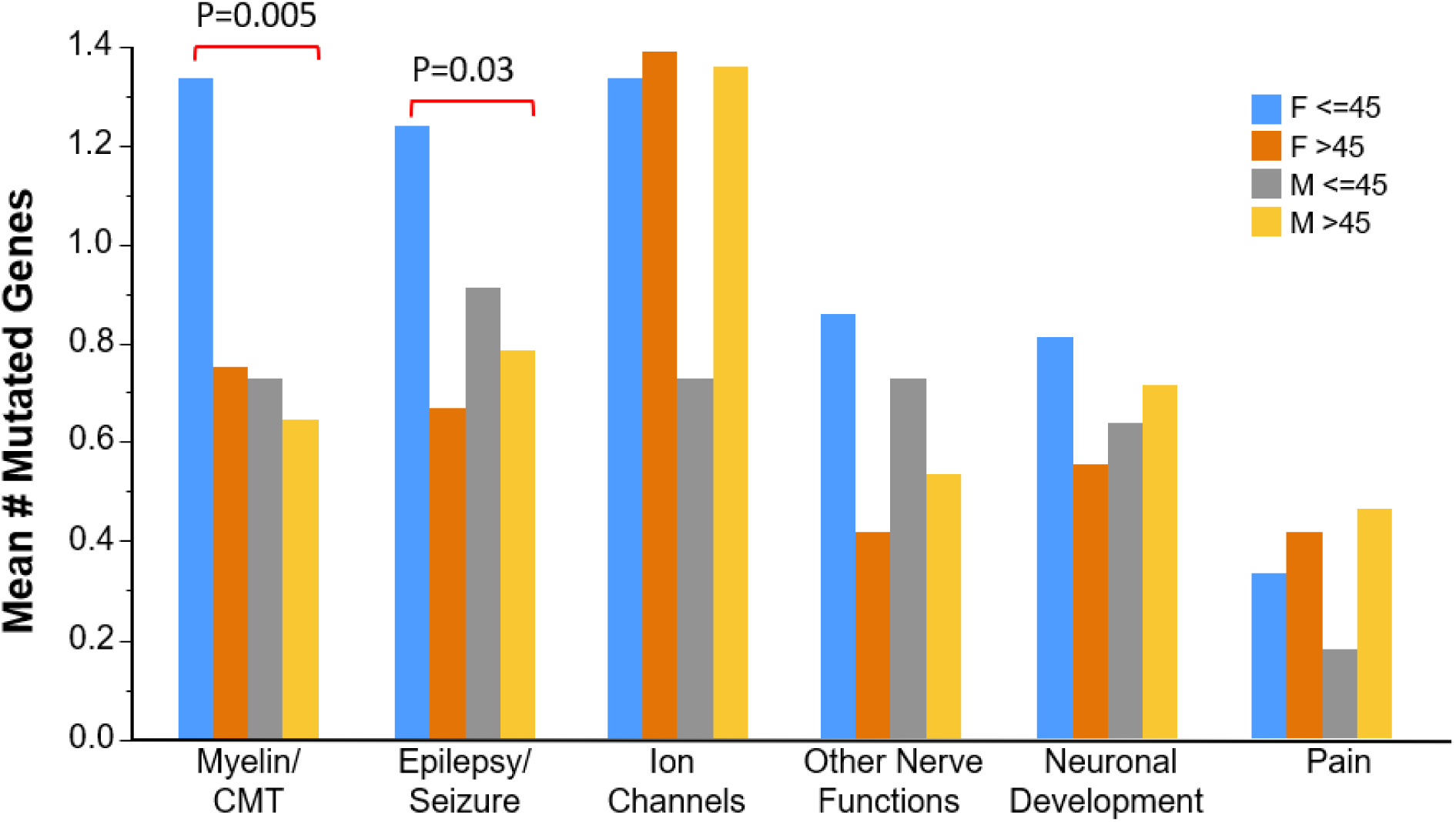
Mean numbers of genes with rare mutations in 100 TN patients. Whole Genome Sequencing (WGS) data with genes prioritized by bioinformatics prediction of mutations’ impact on protein function, low Minor Allele Frequency (MAF) and symptom sharing with single gene disorders. Stratified by sex and age of onset (<=45 v. >45) and by the genes’ known biological function or disease associations.

**Table 2:**
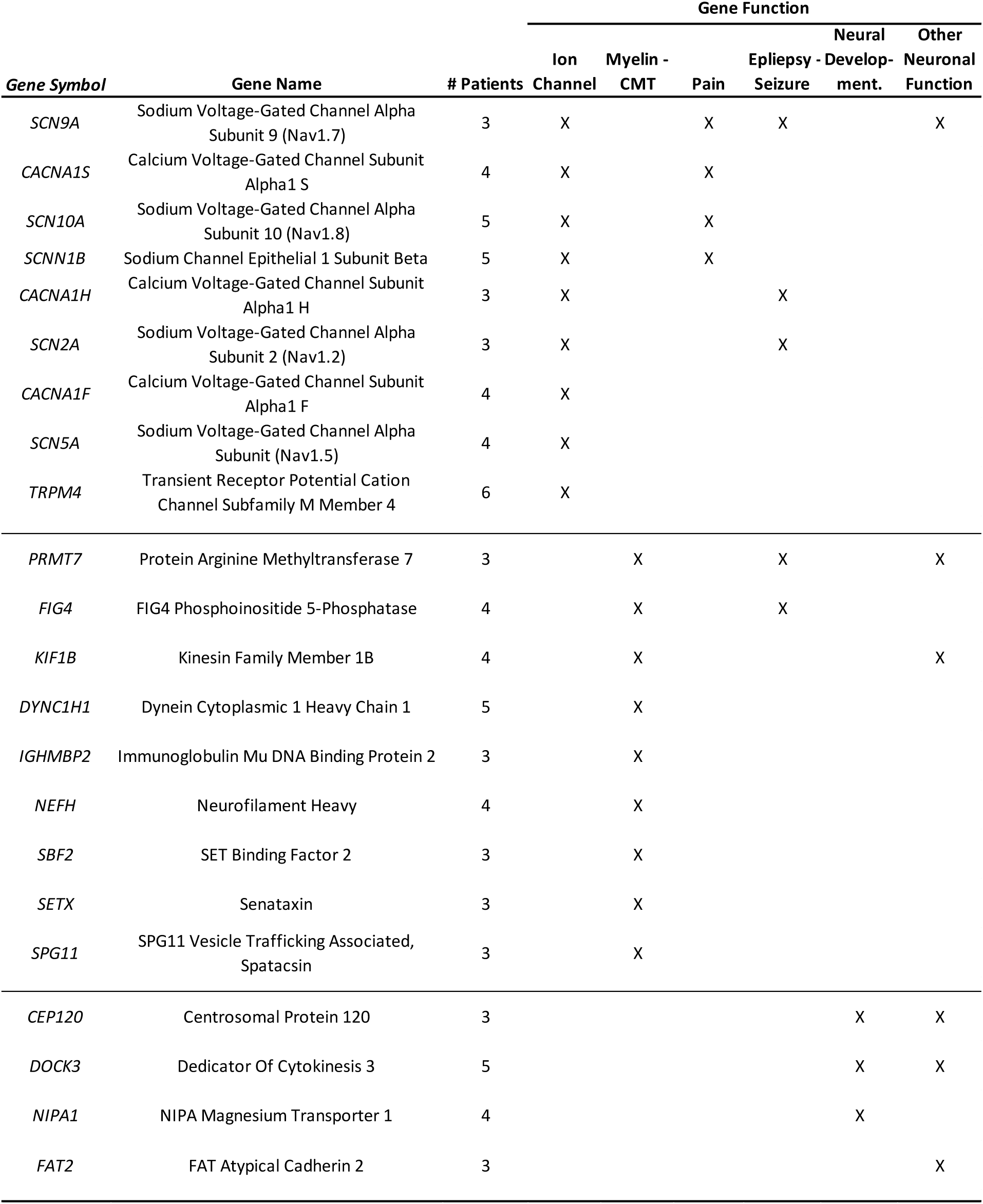
Genes with rare, potentially damaging mutations in three or more TN patients. Includes nine genes coding for ion channels and nine genes coding for myelin-related functions or are associated with Charcot-Marie-Tooth (CMT) disease.

## Discussion

The findings of this study and previously reported genetic research suggest that Trigeminal Neuralgia is a complex, multifactorial disorder. The aetiology of TN1 appears highly heterogeneous, with both common polymorphisms and rare variants contributing to risk. Evidence from this study also suggests that genetic variants may have different effects on risk depending on interactions with patients’ sex, age of onset, and clinical subtype (Classical versus Idiopathic). Data regarding these variables are not usually available for patients participating in the large biobank cohorts. Going forward, obtaining more comprehensive phenotypic assessment data of TN patients participating in large scale biobanks, where feasible, would be very valuable.

This study’s GWAS and WGS findings identify numerous genes with known functions in pathways relevant to the pathophysiology of TN. For example, the Potassium Two Pore Domain Channel Subfamily K Member 10 gene (*KCNK10*) suggested by our GWAS acts as an open rectifier ion channel which primarily passes outward current under physiological K+ concentrations (Stelzer et al., 2016). It appears to play a role in the excitability of trigeminal ganglion neurons (Chang et al., 2021) and its expression in axotomized rat primary sensory neurons reduced neuropathic pain behavior (Messina et al., 2022). Our evidence suggests that that SNPs located >145 kbp from the *LRP1B* gene are strongly association with TN1. This gene encodes for the LDL Receptor Related Protein 1B. However, at this distance from the gene, it is unclear whether these variants influence the gene’s function. Of note, it has been reported that this gene has potential for development of a novel approach to the treatment of chronic pain (García-Fernández et al., 2021).

This study has identified significant associations between TN1 and genes not previously considered candidates for TN nor any other form of pain, acute or chronic. For example, the Cell Adhesion Molecule L1 Like (*CHL1*) gene plays a role in nervous system development, nerve regeneration, synaptic plasticity, regulation of the efficacy of GABAergic synapses, and suppresses neuronal cell death and may be important for nerve regeneration (Stelzer et al., 2016). A SNP in an intron of the Zinc Finger Protein 804B (*ZNF804B*) gene exhibited association with the surface area of the right Heschl’s gyrus region of the brain in healthy individuals (Cai et al., 2014). The Cut Like Homeobox 1 (*CUX1*) gene has been shown to control synaptogenesis, neuronal differentiation in the brain, regulate dendrite development and branching, as well as dendritic spine formation in the cortex (Stelzer et al., 2016). Finally, the Sphingomyelin Synthase 1 (*SGMS1*) gene was shown to be involved in regulation of membrane KCNQ1/KCNE1 channel density on neuronal membranes (Wu, 2016).

Three previous reports of TN-causing mutations using Whole Genome or Whole Exome Sequencing evaluated 7 (Wang et al., 2023), 11 (DiStefano et al., 2020) and 290 (Dong et al., 2020) TN patients, and reported finding mutations in 1, 36 and 82 genes, respectively. The first study focused on the *MARS1* gene and the search for mutations in the second study was limited to a panel comprised of 173 ion channel genes encoding sodium, potassium, calcium, chloride, transient receptor potential channels, and gap junction channels. The third study of 290 TN cases used several strategies to prioritize genes likely to cause TN including those with *denovo* mutations and genes in pathways identified using Gene Ontology (GO) enrichment analysis (Dong et al., 2020).

Only one gene, the Sodium Voltage-Gated Channel Alpha Subunit 5 (SCN5A) that mediates voltage-dependent sodium ion permeability of excitable membranes), was identified in this study and two of the previous sequencing studies (Distefano et al., 2020; Dong et al., 2020) (Supplementary Figures S3 and S4 and Supplementary Table S3). SCN5A is involved in voltage-dependent sodium ion permeability of excitable membranes. It has been linked to cardiomyopathies such as Long QT and Sudden Infant Death Syndromes but has not previously been considered a top candidate for neuropathic pain. This study identified 16 genes also reported by DiStefano et al. (2020), 7 genes reported by Dong et al. (2020) and the *MARS1* gene identified by Wang et al. (2023). Combining previously reported mutations with findings from this study, a total of 510 mutations (490 unique and 20 duplicate pairs) were found in 277 unique genes (Supplementary Table S5).

Phevor scores used to rank mutations based on HPO terms labelled Models 2 and 3 (Supplementary Table S3) are highly correlated (r = 0.99) but both models are only moderately correlated with Model 1 that has only the one HPO term HP: 0100661 Trigeminal Neuralgia (r = 0.58 and 0.60). This indicates that evaluations performed under different HPO models can yield substantially different results so exploration of multiple HPO models should be considered in future studies using the symptom sharing approach for identifying candidate genes.

We hypothesized that the aetiology of TN may differ between patients who primarily experience severe paroxysmal pain (TN1) versus patients who report substantial concomitant constant orofacial pain in addition to paroxysmal pain (TN2). Previously reported GWAS analyses of TN patients based on data from biobanks (Dong et al., 2020; Hale et al., 2021) revealed significant associations for SNPs on chromosomes 1 and 5 in one study (Hale et al., 2021) but these were not replicated in the present study, nor were any of the findings of the present study observed in the previous one. Cases used for the biobank association studies included both TN patients with (TN2) and without (TN1) substantial concomitant constant pain. If some genetic associations are different in TN1 versus TN2 subtypes, a case group made up of a mixture of these subtypes may produce different results than our study where cases with significant constant orofacial pain were excluded. Although we are unaware of any direct tests of differences in genetic associations between the two subtypes, TN1 and TN2 patients exhibited striking differences in analgesic response in a clinical trial of a monoclonal antibody against the calcitonin gene-related peptide (CGRP) receptor (Schott Andersen et al., 2022). TN1 patients responded positively to the drug more often than to placebo, while TN2 patients responded to the drug less often than placebo (Fisher’s Exact Test P-value 0.039, Supplementary Figure S5). While this small study needs to be replicated, it is consistent with our hypothesis that these TN subtypes may have differences in aetiology that should be considered in both future research studies and clinical trials.

Limitations of the study include its modest samples size. However, our strategy of focusing recruitment exclusively on patients experiencing only severe paroxysmal pain (TN1) without substantial concomitant constant pain (TN2) may make our modest sized sample more homogeneous and thus indirectly increase statistical power for detecting aetiologically relevant genetic variants. The other major limitation is lack of detailed information regarding neurovascular compression of the trigeminal nerve. This information was available for only 299 of our study’s 626 TN1 patients, and for many of these the available information only consists of a binary variable of “having/not having” compression. Severity of compression can range from blood vessels barely touching the nerve and thus unlikely to have caused injury to frank compression that is far more likely to be damaging (Maarbjerg et al., 2015; Brînzeu et al., 2018). Severe compression may be associated with genetic variants related to myelin and repair of neuronal damage, as well as neuronal hyperexcitability triggered by axonal injury. Blinded, and standardized assessment based on MRI imaging, as well as video and case notes for those undergoing surgery, where available, will be invaluable for further investigations of the role that NVC plays in the aetiology of TN, including potential interactions with genetic and other important risk factors.

## Data Availability

Summary statistics and a deidentified phenotype file will be provided upon request to SRD at. Patients complete individual WGS and GWAS profiles are confidential and cannot be released. Contribution of the findings to collaborative consortium efforts may be permissible with adequate protections of subject privacy.

## Contributors

KJB, ZS and SRD designed the study. SRD wrote the manuscript. SRD, OAK, FK and C-YH accessed and verified the data, carried out the statistical analysis, and created the figures. All authors were involved in data collection, data interpretation, and drafting of the manuscript. All authors critically reviewed the manuscript and approved the final version.

## Declaration of interests

KJB reports grants from the Facial Pain Research Foundation, and personal fees from ClearPoint Neuro not related to this research. OAK reports non-grant support from Rutgers School of Dental Medicine. ZS reports grants from the Facial Pain Research Foundation for support of this study, and from Algogene Pain Genetics and Pfizer Neuropathic Pain Foundation for research outside the work submitted here. SRD reports grants from the Facial Pain Research Foundation, the Foundation of the University of Medicine and Dentistry of New Jersey, ongoing support of Rutgers School of Dental Medicine for this study and personal fees from Kriya Therapeutics. All other authors declare no competing interests.

## Data sharing

Summary statistics and a deidentified phenotype file will be provided upon request to SRD at. Patients’ complete individual WGS and GWAS profiles are confidential and cannot be released. Contribution of the findings to collaborative consortium efforts may be permissible with adequate protections of subjects’ privacy.

## Acknowledgments

This study was funded by grants from the William H. and Leila A. Cilker Genetics Research Program of the Facial Pain Research Foundation, the Foundation of the University of Medicine and Dentistry of New Jersey, and ongoing support of Rutgers School of Dental Medicine. This study used the high-performance computational resources of the Amarel LINUX cluster created and supported by Rutgers’ Office of Advanced Research Computing (OARC). The following individuals played leading roles in patient recruitment and assessment at their respective clinical sites:

- *Andrew H Ahn, M.D.* (University of Florida, College of Medicine, Department of Neurology, Gainesville, FL)
- *John Alksne, M.D.* (University of California, School of Medicine, Department of Neurological Surgery, La Jolla, CA)
- *Edward F Chang, M.D.* (University of California San Francisco, School of Medicine, Department of Neurological Surgery, San Francisco, CA)
- *Mojgan Hodaie, M.D., M.Sc.* (University of Toronto, Faculty of Medicine, Department of Surgery, Toronto, Canada)
- *John YK Lee, M.D.* (University of Pennsylvania, Perelman School of Medicine, Department of Neurosurgery, Philadelphia, PA)
- *Shirley McCartney, Ph.D.* and *Samantha Yau, B.S.* (Oregon Health & Science University, School of Medicine, Department of Neurological Surgery, Portland, OR)
- *Konstantin Slavin, M.D.* (University of Illinois, College of Medicine, Department of Neurosurgery, Chicago, IL)
- *Joanna M Zakrzewska* B.D.S., M.B. B.Chir., M.D. (Pain Management Centre, National Hospital for Neurology & Neurosurgery, London, UK)

The contributions of subjects who volunteered to participate in the study are also recognized and appreciated.

## Supplementary Figures

**Figure S1:**
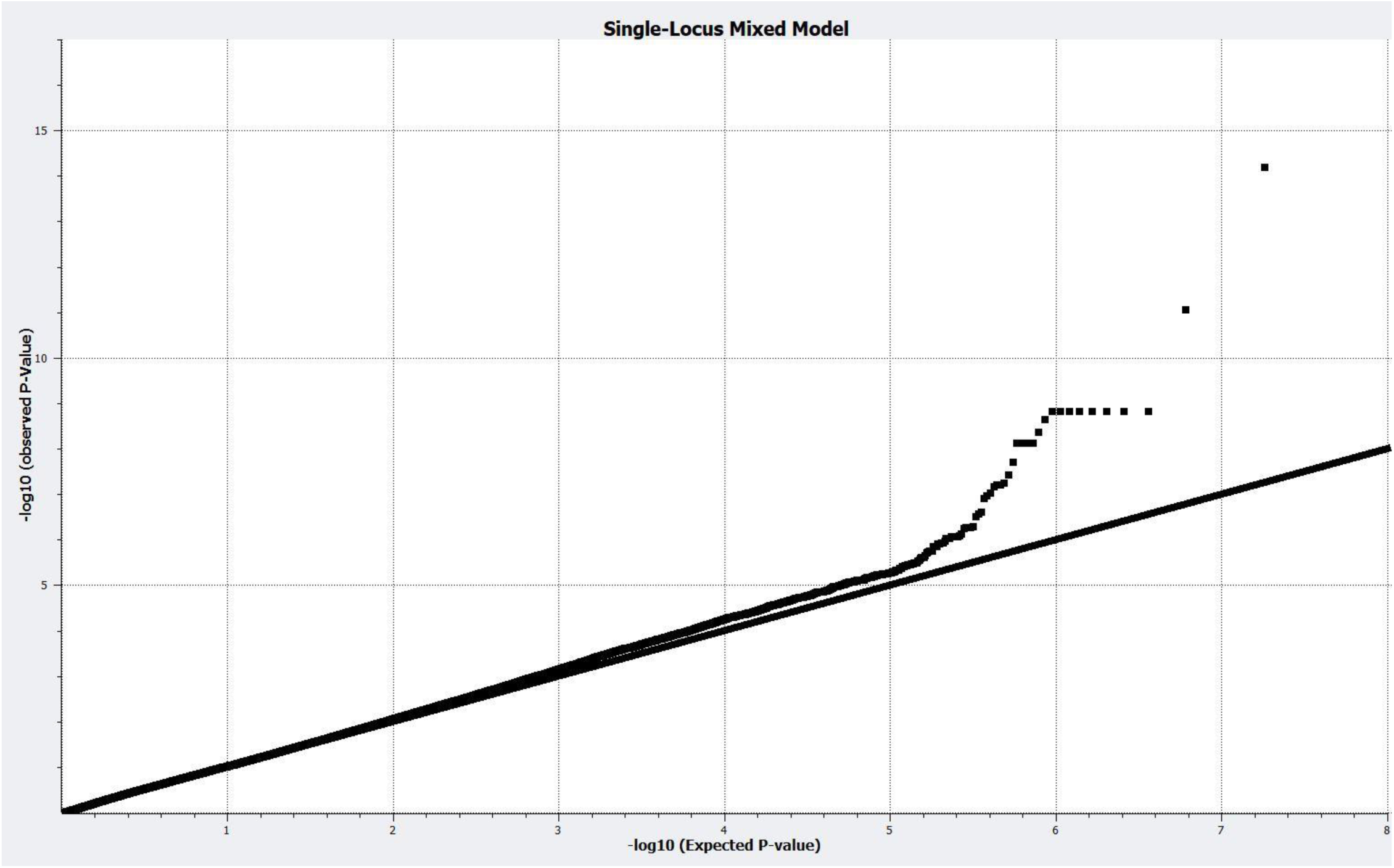
Single Locus Mixed Model Avoids P-value Inflation. Q-Q Plot of Observed versus Expected -Log10 P-values for Mixed Model Additive Analysis.

**Figure S2:**
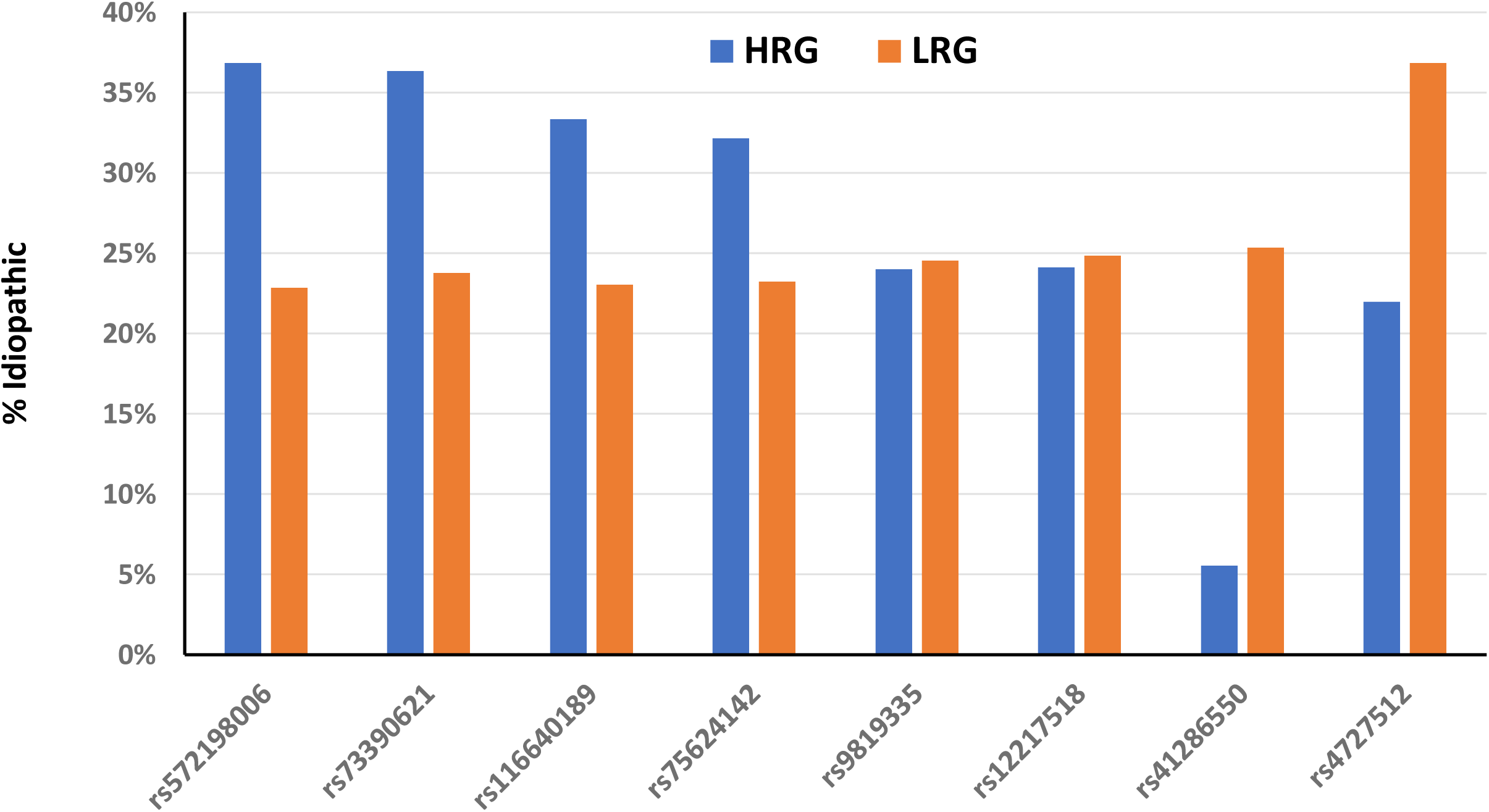
Percentage of TN patients Idiopathic by HRG v. LRG for 8 GWAS SNPs. Patients who inherited the HRG group are more often Idiopathic in the four SNPs on the left, of approximately equal frequency for two SNPs in the middle. The Idiopathic frequency of the HRG group (6%) is lower than the LRG group (25%) at SNPs rs41286550 located in the *KCNK10* gene (P-value 0.084). The Idiopathic frequency of the HRG group (22%) is lower than the LRG group 37%) at SNPs rs4727512 located in the *CUX1* gene (P-value 0.063).

**Figure S3:**
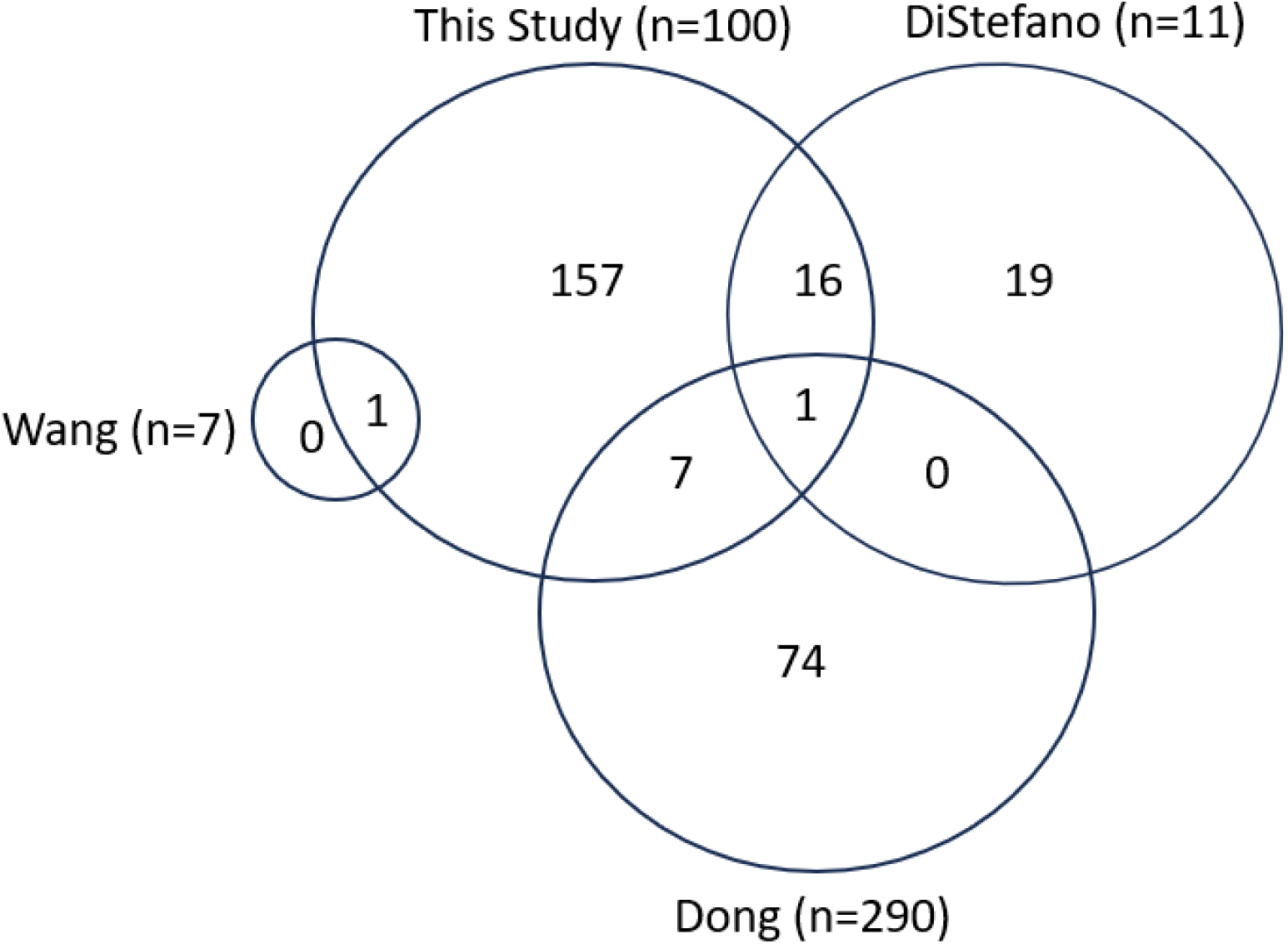
Overlap of Genes with Rare Candidate Mutations Across Four Studies. Only 1gene (*SCN5A*) has been identified in this study and two previous studies (n = numbers of patients sequenced in each study). As shown *in Figure S6*, seven other genes were identified in both this study and the study reported by Dong et al. (2020) including *CACNA1H* and 16 genes overlap between this study and the study reported by DiStefano et al. (2020) including *SCN9A, KCNC3* and *CACNA1l*.

**Figure S4:**
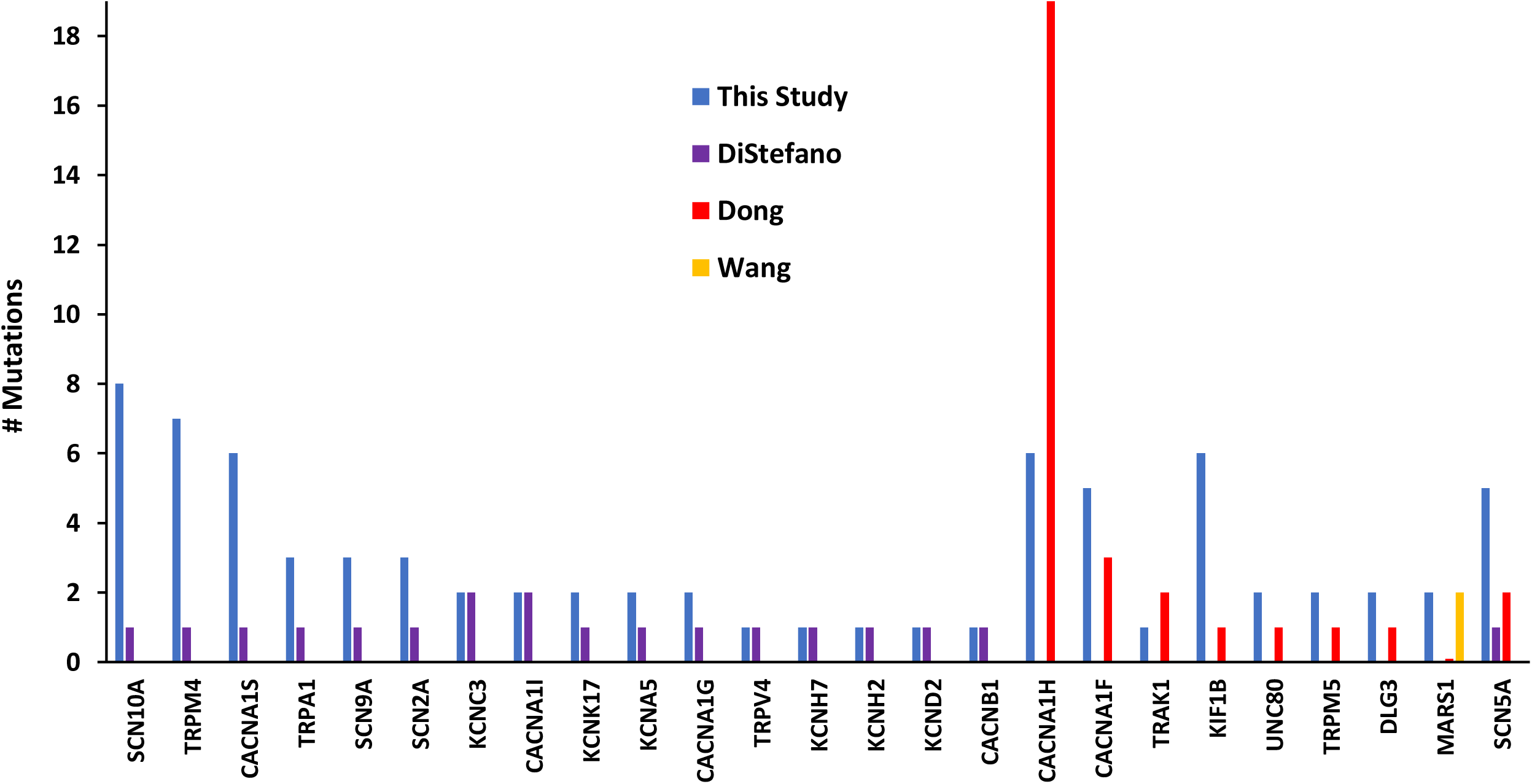
TN Candidate Genes with Rare Mutations Identified in Multiple Studies. Whole Genome and Whole Exome Sequencing studies identified a total of 301 genes with rare mutations predicted to alter protein function in pathways related to the biology of TN.

**Figure S5.**
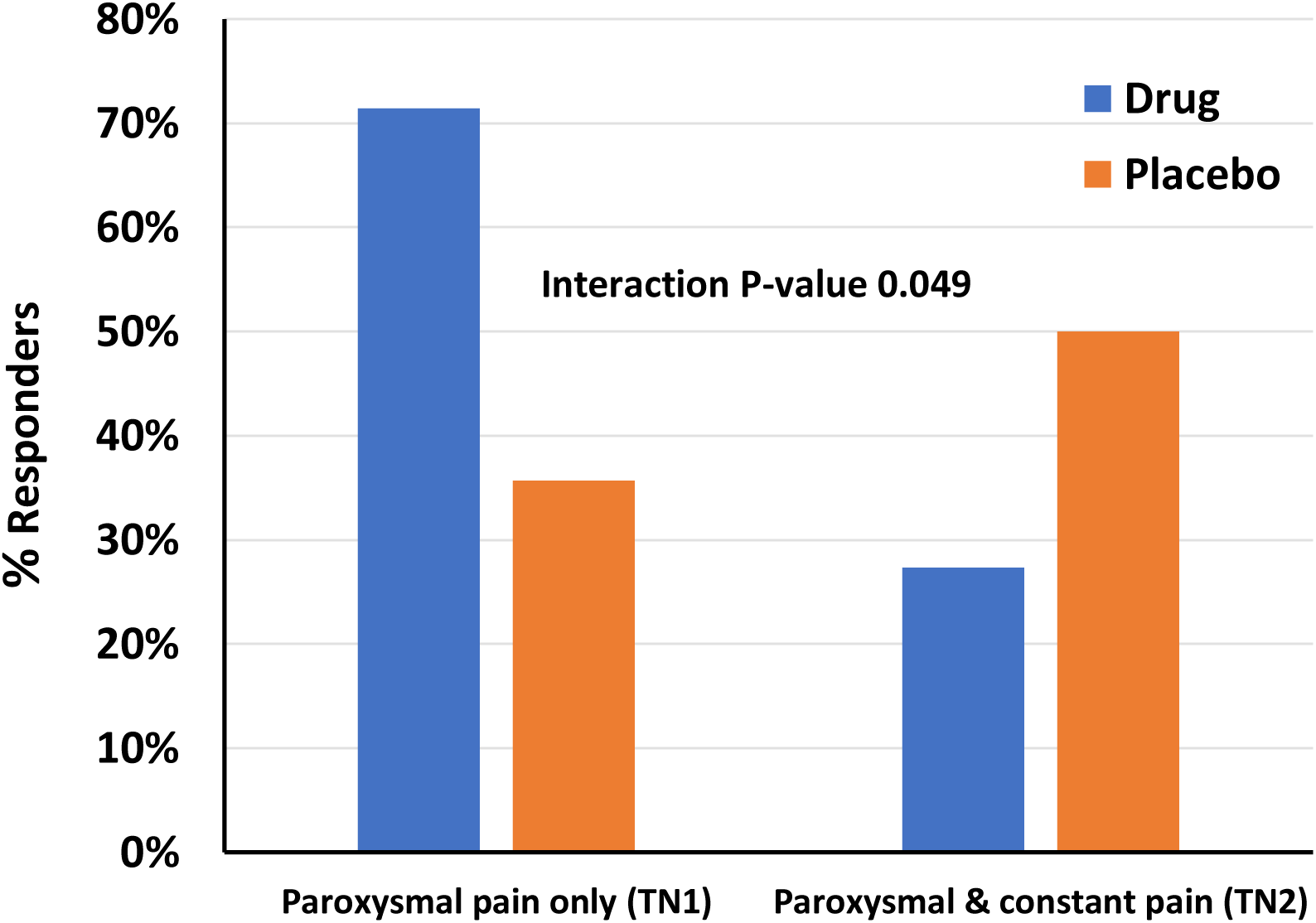
TN Subtypes’ Responses to CCRP Receptor Antibody. 71% of TN patients with paroxysmal pain only (TN1) respond to Erenumab antibody against the calcitonin gene-related peptide (CGRP) receptor (higher frequency than their placebo group) while only 27% of TN patients reporting both paroxysmal and constant pain (TN2) respond (lower than their placebo group). Fisher’s Exact Test P-value = 0.039 [data from: Schott Andersen et al., 2022].

## Supplementary Tables

**Table S1:** Study Demographics by Recruitment Site. Summary for all subjects included in the GWAS analysis.

| Site | # Patients | % Female | Female Mean AOO* | Female AOO Range | Male Mean AOO | Male AOO Range |
| --- | --- | --- | --- | --- | --- | --- |
| Chicago, USA | 15 | 80% | 54.7 | 29 - 85 | 51.3 | 31 - 62 |
| Gainesville, USA | 117 | 66% | 57.4 | 19 - 90 | 63.3 | 17 - 89 |
| London, UK | 45 | 69% | 57.5 | 34 - 81 | 50.9 | 31-63 |
| New York, USA | 18 | 56% | 59.8 | 30 - 86 | 63.6 | 42 - 87 |
| Philadelphia, USA | 73 | 68% | 53.1 | 18 - 80 | 51.2 | 29 - 79 |
| Portland, USA | 253 | 66% | 53.6 | 15 - 91 | 54.0 | 17 - 83 |
| San Diego, USA | 12 | 58% | 58.2 | 28 - 73 | 53.0 | 43 - 65 |
| San Francisco, USA | 34 | 59% | 55.1 | 19 - 77 | 62.1 | 40 - 81 |
| Toronto, CA | 59 | 56% | 53.2 | 21 - 86 | 54.7 | 23 - 78 |
| **All Cases** | **626** | **65%** | **54.9** | **15 - 91** | **56.1** | **17 - 89** |
| **Controls** | **827** | **57%** | **n/a** | **n/a** | **n/a** | **n/a** |

\*Age Of Onset

**Table S2:**
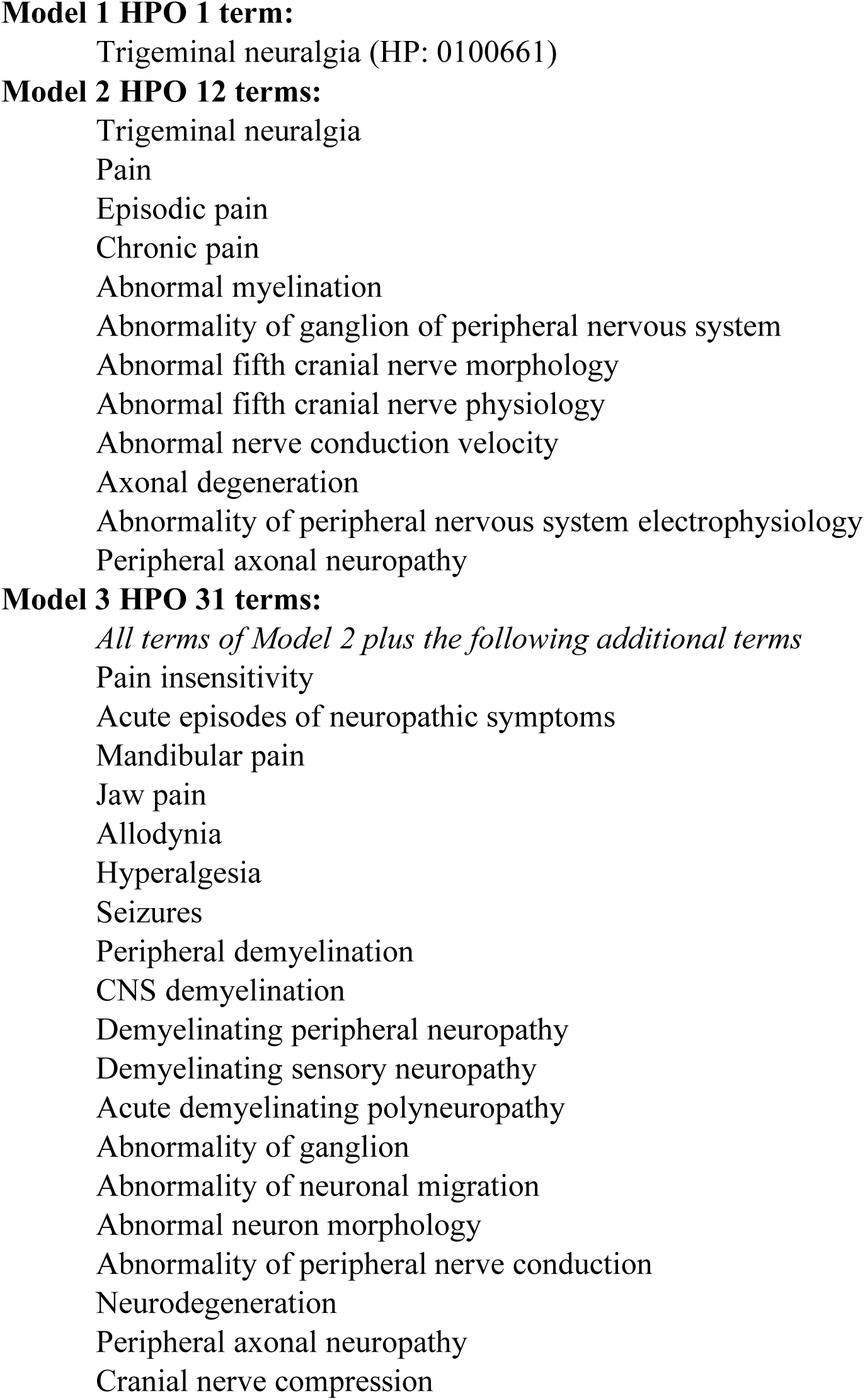
Human Phenotype Ontology (HPO) Terms Used for NGS Search. Standardized vocabulary of phenotypic abnormalities encountered in human disease. Three models analyzed for the study ranged from the very narrow Model 1 to broader Models 2 and 3. The following Excel files will be provided as email attachments upon request to

***Table S3: Candidate mutations discovered in 100 TN1 Cases***

349 candidate top ranked mutations for TN1 based on Fabric Genomics’ Phevor Scores with eQTL and sQTL associations where available.

Excel File: TN_GeneticsStudy_349Mutations_TableS3_10July2024

***Table S4: Candidate Genes discovered in 100 TN1 Cases***

182 genes, biological functions, disease associations, # mutations & # patients affected. Excel File: TN_GeneticsStudy_182Genes_TableS4_10July2024

***Table S5: Candidate Mutations: This Study & Previous Reports***

510 mutations (490 unique) in 277 genes discovered in this study and studies reported previously by DiStefano et al. (2020), Dong et al. (2020) and Wang et al. (2023).

Excel file: TN_Genetic_Studies 510Mutations_All Studies_10July2024

## Notes

### Author Declarations

Ethical review and approval by the institutional review board (IRB) of Oregon Health & Science University

